# Automated mood disorder symptoms monitoring from multivariate time-series sensory data: Getting the full picture beyond a single number

**DOI:** 10.1101/2023.03.25.23287744

**Authors:** Filippo Corponi, Bryan M. Li, Gerard Anmella, Ariadna Mas, Miriam Sanabra, Eduard Vieta, INTREPIBD Group, Stephen M. Lawrie, Heather C. Whalley, Diego Hidalgo-Mazzei, Antonio Vergari

## Abstract

Mood disorders are among the leading causes of disease burden worldwide. They manifest with changes in mood, sleep, and motor-activity, observable with physiological data. Despite effective treatments being available, limited specialized care availability is a major bottleneck, hindering preemptive interventions. Near-continuous and passive collection of physiological data from wearables in daily life, analyzable with machine learning, could mitigate this problem, bringing mood disorders monitoring outside the doctor’s office. Previous works attempted predicting a single label, e.g. disease state or a psychometric scale total score. However, clinical practice suggests that the same label can underlie different symptom profiles, requiring personalized treatment. In this work we address this limitation by proposing a new task: inferring all items from the Hamilton Depression Rating Scale (HDRS) and the Young Mania Rating Scale (YMRS), the most-widely used standardized questionnaires for assessing depression and mania symptoms respectively, the two polarities of mood disorders. Using a naturalistic, single-center cohort of patients with a mood disorder (N=75), we develop an artificial neural network (ANN) that inputs physiological data from a wearable device and scores patients on HDRS and YMRS in moderate agreement (quadratic Cohen’s *κ* = 0.609) with assessments by a clinician. We also show that, when using as input physiological data recorded further away from when HDRS and YMRS were collected by the clinician, the ANN performance deteriorates, pointing to a distribution shift, likely across both psychometric scales and physiological data. This suggests the task is challenging and research into domain-adaptation should be prioritized towards real-world implementations.

## 1 Introduction

Mood disorders, also referred to as affective disorders, are a group of diagnoses in the Diagnostic and Statistical Manual 5^th^ edition (DSM-5) [3] classification system. They are a leading cause of disability worldwide and a major contributor to the overall global burden of disease [63], with an estimated total economic cost greater than USD326.2 billion in the United States alone [31]. They encompass a variety of symptom combinations affecting mood, motor activity, sleep, and cognition and manifest in episodes categorized as major depressive episodes (MDEs), featuring feelings of sadness and loss of interest, or, at the opposite extreme, (hypo)manic episodes (MEs), with increased activity and self-esteem, reduced need for sleep, expansive mood and behavior. According to the DSM-5 classification system, MDEs straddle two nosographic constructs, i.e. Major Depressive Disorder (MDD) and Bipolar Disorder (BD), whereas MEs are the earmark of BD only [80].

Clinical trials to this day entirely rely on clinician-administered standardized questionnaires for assessing mood disorders symptoms severity. However, the psychiatric community has increasingly been looking to biomarkers that could help validate or push forward the current understanding of mental disorders, providing some measurable and objective indicators [15]. Hamilton Depression Rating Scale-17 (HDRS) [32] and Young Mania Rating Scale (YMRS) [83] are the two most-widely used clinician-administered scales for depressive and manic symptoms respectively; they are commonly used for setting treatment outcome criteria and tracking disease course [76]. While compressing these scales to the total sum across their items has been a typical approach in clinical trials [14, 22], a focus on individual symptoms allows for a richer clinical description. For instance, considering individual items can identify drug specificity for symptom domains, thereby enabling tailored treatment [49, 81]. This modus operandi is also found in everyday psychiatric practice where the specialist, when recommending a given intervention, takes into account the specific features of a patient, including their symptom patterns, beyond a reductionist disease label [62, 68]. Figure 1 shows the HDRS and YMRS items and illustrates how patients sharing the same nosographic category and the same severity level (defined from binning HDRS and YMRS total score as in Tohen et al. [77]) can “look” clinically different. The HDRS and YMRS full questionnaires are provided in Appendix B and Appendix C for reader’s convenience.

**Figure 1:**
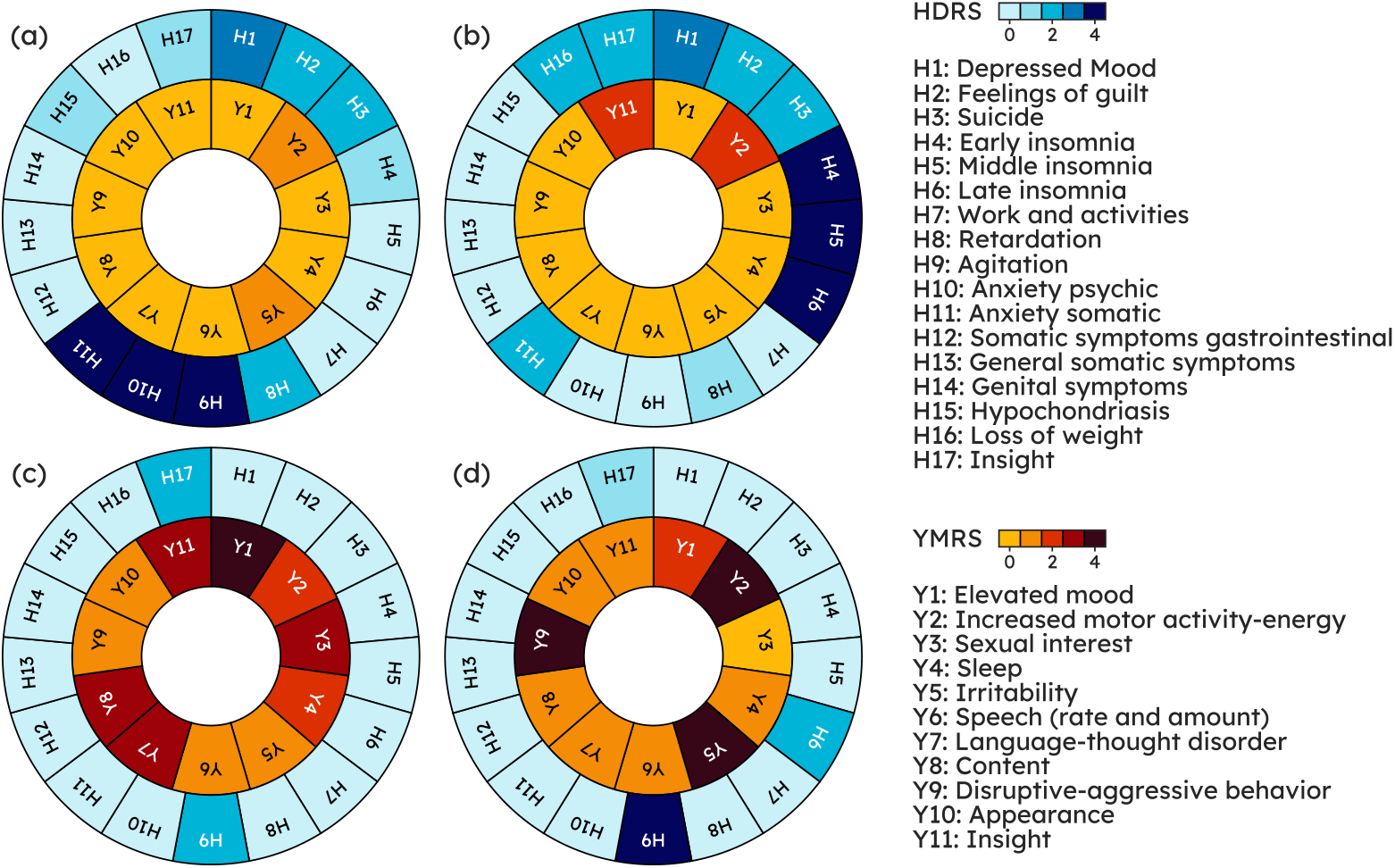
Patients sharing diagnostic category and severity level can have different clinical profiles. Top row shows a pair of patients with Major Depressive Disorder (MDD) on a depressive episode. While both share the same severity levels, total HDRS ≥ 23 [77], patient (a), with HDRS totaling 24, exhibits high levels of anxiety (*H9, H10, H11*), whereas patient (b), with HDRS totaling 26, displays a marked insomnia component (*H4, H5, H6*). Bottom row shows a pair of patients with Bipolar Disorder (BD) on a manic episode with total YMRS ≥ 25 [77]. Patient (c), with YMRS totaling 30, has an irritable/aggressive profile (*Y2, Y5, Y9*) whereas patient (d), with YMRS totaling 30, has a prominently elated/expansive presentation (*Y1, Y3, Y7, Y11*). The two pairs show that the same severity level and the same (or a similar) total score can be realized from different combinations of symptoms, giving rise to different clinical presentations that may benefit from bespoke treatment and management. HDRS and YMRS items and normalized scales are displayed on the right-hand side of the figure and the full scales are available for reader’s convenience in Appendix B and Appendix C.

Low availability of specialized care for mood disorders, with rising demand straining current capacity [59, 65], is a major barrier to symptom monitoring, resulting in long waits for appointments and reduced scope for pre-emptive interventions. Furthermore, psychiatric interviews only sample a snapshot of the clinical course and recall bias can affect patients’ report of their conditions in the time between appointments [35]. The fortunate conjuncture of advances in machine learning (ML) and, on the other hand, widespread adoption in the general population of increasingly miniaturized and powerful wearable devices set the stage for personal sensing [52]. This involves near-continuous and passive collection of data from sensors embedded in the context of daily life, with the aim of identifying digital biomarkers associated with mental health symptoms at the individual level. Personal sensing promises to lower barriers to healthcare access and shift the healthcare paradigm from reactive to proactive. Mood disorders stand out as particularly amendable to modeling with digital biomarkers since their predominant features (disturbances in mood, sleep patterns, and motor activity) correlate with changes in physiological parameters, conveniently measurable with wearable sensors. Indeed, changes in the autonomic system, as captured with electrodermal activity, heart rate variability, as well as actigraphy patterns (instances of digital biomarkers) have been linked to mood-states [27, 64, 74].

Studies aiming to automate psychological states detection have been burgeoning on the back of this unprecedented opportunity. However, most examined non-clinical phenotypes (e.g. stress) and recruited general populations (e.g. college students) were assessed with self-reported questionnaires and commercial wearable devices [6, 69, 78]. Only few previous endeavors specifically addressed mood disorders using clinician assessments. Côté-Allard et al. [12] explored a binary classification task, that is distinguishing subjects with BD on a ME from different subjects with BD recruited outside of a disease episode, when stable. The study experimented with different subsets of pre-designed features from wristband data and proposed a pipeline leveraging features extracted from both short and long segments taken within 20-hour sequences. Pedrelli et al. [57], expanding on Ghandeharioun et al. [30], used pre-designed features from a wristband and a smartphone to infer HDRS residualized total score (derived by subtracting baseline total score from total score at following assessments) with traditional ML models. Tazawa et al. [73] employed gradient boosting with pre-designed features from wristband data and pursued case-control detection in MDD and, secondarily, HDRS total score prediction. Similarly, a brief communication by Jacobson et al. [36] predicted case-control status in MDD from actigraphy features with gradient boosting. Nguyen et al. [53] used a sample including patients with either schizophrenia or MDD wearing an actigraphic device and explored case-control detection where schizophrenia and MDD were either considered jointly (binary classification) or as separate classes (multi-class classification). Of notice, this was the first work to apply artificial neural networks (ANNs) directly on minimally processed data, showing that they outperformed traditional machine learning models. Lastly, the work of Lee et al. [43], using a large nationwide South Korean cohort of patients with a mood disorder, including both MDD and BD, investigated mood episode prediction with a random forest and derived features from wearable and smartphone data. Table 1 reports the works cited above and shows how they all pursued either binary/multi-class classification or HDRS (residualized) total score regression. All but one extracted pre-designed features from wearable data. Finally, it should be noted that, since collecting data from patients on an acute episode, using specialist assessments and research-grade wearables, is a challenging and expensive enterprise, the sample size of previous studies typically range from tens of patients to a few dozens (with the exception of Lee et al. [43] with 270 patients).

**Table 1:** Our work is the first attempting to infer the full mood disorders symptom profile from physiological wearable data, providing actionable clinical information beyond a single reductionist label. Previous studies recruiting patients with either a Diagnostic and Statistical Manual of Mental Disorders (DSM) or an International Classification of Diseases (ICD) mood disorder diagnosis, using psychiatrist assessments and passively collected wearable data are herewith reported. BD: Bipolar Disorder; F_%_: Percent Females; M_age_: mean age; MAE: Mean Absolute Error; MDD: Major Depressive Disorder; SCZ: Schizophrenia; SD_age_: standard deviation age

|  | DEVICE(S) | NUM.<br>PATIENTS | PATIENTS<br>FEATURES | TASK |
| --- | --- | --- | --- | --- |
| **THIS WORK** | EMPATICA E4 | 75 | MDD, BD; $M_{AGE}=44.16$<br>$SD_{AGE}=14.42$ $F_{\%}=56$ | HDRS AND YMRS ITEMS<br>MULTI-TASK REGRESSION |
| [12] | EMPATICA E4 | 47 | BD; $M_{AGE}=44$<br>$SD_{AGE}=15$ $F_{\%}=67.24$ | MANIA VS EUTHYMIA<br>BINARY CLASSIFICATION |
| [30] | EMPATICA E4<br>AND ANDROID PHONE | 12 | MDD; $M_{AGE}=37$<br>$SD_{AGE}=17$ $F_{\%}=75$ | HDRS TOTAL SCORE<br>REGRESSION |
| [57] | EMPATICA E4<br>AND SMARTPHONE | 31 | MDD; $M_{AGE}=33.7$<br>$SD_{AGE}=14$ $F_{\%}=74$ | HDRS TOTAL SCORE<br>REGRESSION |
| [36] | ACTIWATCH | 23 | MDD; $M_{AGE}=48.2$<br>$SD_{AGE}=11.0$ $F_{\%}=43$ | DEPRESSION DETECTION<br>BINARY CLASSIFICATION |
| [73] | SILMEE W20 | 45 | MDD, BD; $M_{AGE}=52.1$<br>$SD_{AGE}=13.2$ $F_{\%}=46.7$ | DEPRESSION DETECTION<br>BINARY CLASSIFICATION |
| [53] | ACTIWATCH | 45 | MDD, SCZ; $M_{AGE}=44.70$<br>$SD_{AGE}=11$ $F_{\%}=73.33$ | DISEASE DETECTION<br>BINARY/MULTI-CLASS CLASSIFICATION |
| [43] | FITBIT CHARGE HR 2 OR 3<br>AND SMARTPHONE | 270 | MDD, BD; $M_{AGE}=23.3$<br>$SD_{AGE}=3.63$ $F_{\%}=54.4$ | MOOD EPISODE PREDICTION<br>BINARY CLASSIFICATION |

The contribution of this work is two-fold: **(1)** Taking one step beyond the prediction of a single label, which misses on actionable clinical information towards personalizing treatment, we propose a new task in the context of mood disorders monitoring with physiological data from wearables: inferring all items in HDRS (17 items) and YMRS (11 items), as scored by a psychiatrist. The device we used was the Empatica E4 wristband [24] which monitors acceleration, blood volume pressure, heart rate, inter-beat intervals, electrodermal activity, and skin temperature, with no active interaction required of the user. **(2)** We investigate some of the methodological **challenges** associated with the task at hand and explore possible ML solutions. **c1**: inferring multiple (28) target variables, i.e. multi-task learning (MTL). **c2**: modeling ordinal data, such are HDRS and YMRS items. **c3**: learning subject invariant representations, since, especially with noisy data and a sample size in the order of dozens, models tend to focus on subject specific features to master the task at hand leading to poor generalization [54]. **c4**: learning from imbalance data, as patients on a acute episode usually receive intensive treatment and acute states therefore tend to be relatively short periods in the overall disease course [17, 79] thereby tilting scores towards low values.

## 2 Results

### 2.1 Study sample & pipeline

The following analyses are based on 75 subjects with a DSM-5 mood disorder diagnosis (either MDD or BD). Subjects recruited on an acute episode had up to four assessments, at different stages of the disease course: **T0** acute phase (upon hospital admission or at the home treatment unit), **T1** response onset (50% reduction in total HDRS/YMRS), **T2** remission (total HDRS/YMRS ≤7), and **T3** recovery (total HDRS/YMRS continuously ≤ 7 for a period of ≥ 8 weeks). On the other hand, subjects with an historical diagnosis but clinically stable at the moment of study admittance (euthymia) were interviewed only once. An overview of the study sample clinical-demographic characteristics is given in Table 3. A total of 149 recording sessions, amounting to over 7000 hours, were available for the present study.

At the start of each assessment, a clinician collected clinical-demographics, including HDRS and YMRS, and provided an Empatica E4 which participants were required to wear on their non-dominant wrist until battery ran out (∼ 48 hours). The wearable records (sampling rate) 3D acceleration (ACC, 32Hz), blood volume pressure (BVP, 64Hz), electrodermal activity (EDA, 4Hz), heart rate (HR, 1Hz), inter-beat intervals (IBI, i.e. the time between two consecutive heart ventricular contractions) and skin temperature (TEMP, 1Hz). IBI was not considered in the present analyses due to extensive sequences of missing values across all recordings, unlike other channels, likely due to high sensitivity to motion and motion artifacts, as suggested previously [67]. Recording sessions were quality-controlled to remove physiologically implausible values, then split into (non-overlapping) segments according to a tunable segment length value *sl* (in real time seconds) and labeled with clinician’s HDRS and YMRS scores. The distribution over item scores was imbalanced towards lower values (Figure A.1), as illustrated by the ratio between the size (i.e. number of segments or equivalently number of recording sessions) of the majority class and of the minority class (referred to *ρ*), which ranged from 4.75 to 99 (median = 20.75).

Some items correspond to symptoms that likely fluctuate over a 48-hour session, especially in an ecological setting where treatments can be administered (e.g., *Y9 disruptive-aggressive behavior* may be sensitive to a sedative drug administered at some point after the beginning of the recording when HDRS and YMRS were scored). To limit this, we isolated segments from the first five hours (**close-to-interview** samples) and used them for the main analysis. Then, in order to study the effect of distribution shift, we tested the trained model on samples from each 30-minute interval following the first five hours of each recording (**far-from-interview** samples). It should be noted that further to a shift in the target variables, a shift in the distribution of physiological data collected with the wearable device is to be expected, owing to different patterns of activity during the day, circadian cycles, and administered drugs. The cut-off of five hours was chosen as a compromise between maximizing samples included in the main analysis and minimizing distribution shift. An illustration of the analysis work-flow is provided in Figure 2.

**Figure 2:**
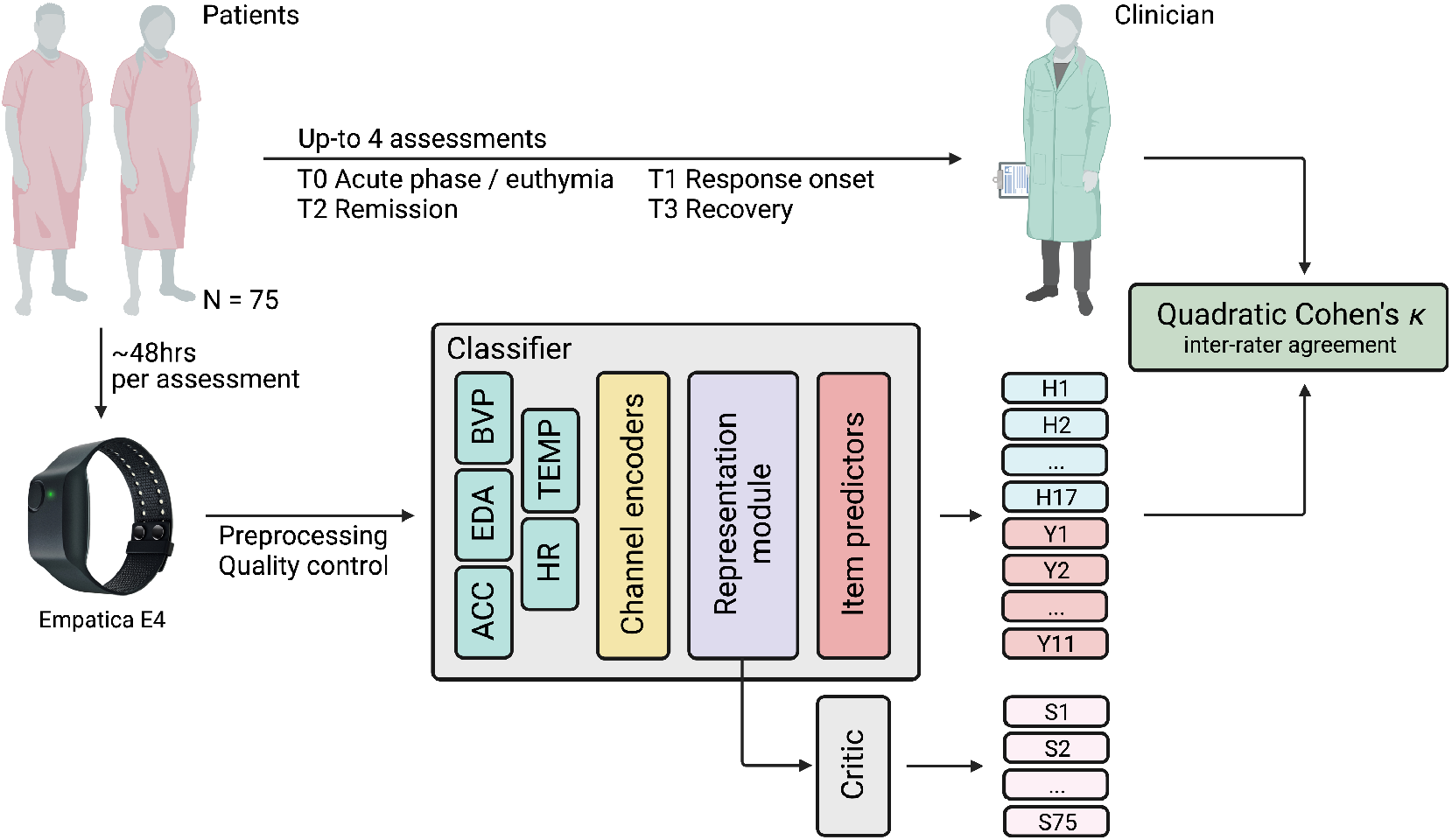
Analysis work-flow. Patients recruited in this study had up to four assessments, corresponding to different disease stages. At the start of each assessment a clinician scored the patient on the Hamilton Depression Rating Scale (H in the figure) and the Young Mania Rating Scale (Y in the figure) and provided an Empatica E4 device asking the patient to wear it for ∼ 48 hours (i.e. average E4 battery life). An ANN model is fed with recording segments and is tasked with recovering the scores issued by the clinician. The quadratic Cohen’s *κ* measures the degree to which the machine scores are in agreement with those from the clinician. The ANN model is made of a classifier CF and a critic CR. The former comprises three main modules: 1) a channel encoder CE, projecting input sensory channels onto a new space where all channels share the same dimensionality, regardless of the native E4 sampling frequency; 2) a representation module RM, extracting a representation *h* that is shared across all items; and 3) one item predictor IP_*j*_ for each item. The critic is tasked with telling subjects (S in the figure) apart using *h* and is pitted in an adversarial game against RM(CE(·)), designed to encourage the latter to extract cross-subjects invariant representations.

### 2.2 Automated Mood Disorder Symptoms Scoring

We used ANNs to model the mapping from physiological recordings to HDRS and YMRS scores. In brief, we had a classifier CF tasked with assigning scores to segments, which comprised three sequential modules: (1) channel encoders (EN) for projecting sensory modalities sampled at different frequencies onto the same space, (2) a representation module (RM) consisting of a single BiLSTM layer for extracting features, and (3) item predictors (IP), as many as there are HDRS and YMRS items, outputting probabilities over scores. This classifier adopted a hard parameter-sharing approach to MTL (**c1**): the channel encoders and the representation module were shared across tasks and extracted a common representation *h* = RM (EN (·)) which was then fed into task-specific item predictors. Losses from different items were then aggregated with a weighted average, to reflect that some items (those with a rank step size of two) count more towards the scale total score. We added a critic CR to the above classifier. CR’s objective was to identify subjects using *h* in an adversarial game, controlled by the hyperparameter *λ*, designed to encourage RM (EN (·)) to extract cross-subjects invariant representations.

As with psychiatry clinical trials, where prospective raters are trained to align with assessments made by an established specialist [2], we used the inter-rater agreement between the ANN and the clinician, measured with the Cohen’s *κ* [51], as evaluation metric for the model. Cohen’s *κ* is a statistic familiar and interpretable to clinicians and clinical trials with multiple rates typically ensure, in the preparation to the study, that a satisfactory level of agreement is reached, in other words that raters converge in the scores they issue. Cohen’s *κ* takes values in [-1, 1], where 1 (−1) means perfect (dis)agreement and a value of 0 indicates random agreement. More specifically, we used the quadratic variant of Cohen’s *κ* (QCK): quadratic weightage penalizes disagreements proportionally to their squared distance, which makes the metric more appropriate to ordinal variables and also less susceptible to imbalanced classes [16, 19, 21, 70]

#### Best model details

On top of the model design described above, we explored a number of ML approaches and discovered the best setting in terms of validation performance through Bayesian Search with Asynchronous HyperBand [46]. We also computed which hyperparameters were the best predictors of the validation QCK. This was obtained by training a random forest with the hyperparameters as inputs and the metric as the target output and deriving the feature importance values for the random forest. The loss type was the hyperparameter most predictive of validation QCK. Table A.1 reports importance values for all hyperparameters. The selected model employed the Cohen’s *κ* loss with quadratic weightage [18] (**c2**), which was preferred to other losses that either treated scores as purely nominal variables or incorporated some notion of orders among ranks. The best model used a (small) critic penalty (*λ* = 0.07) added to the main objective, i.e. scoring HDRS/YMRS. Such a penalty was designed to make CF less reliant on subject-specific information for the main objective and worked by encouraging RM (EN (·)) to conceal subject-specific information from the representation provided to CR (**c3**). However, the training curve showed that the reduction in the main classification loss across epochs was paralleled by the reduction in the (cross-entropy) loss paid by CR, tasked with telling subjects apart. Resampling and loss re-weighting (**c4**) was chosen as a strategy to address class imbalance over other approaches that either re-scale the loss proportionally to the probability assigned to correct rank or re-scale the predicted probabilities by the ranks frequency in the training set. Finally, a segment length *sl* of 16 seconds was selected as best value for segmentation. The highest validation QCK for other choices of *sl* (in seconds) was 0.5167 (8s), 0.5084 (32s), 0.418 (64s), 0.3632 (128s), 0.2543 (256s), 0.1659 (512s), 0.03199 (1024s). Note that *sl* was explored among powers of 2 for computational convenience and that, when segmenting the first 5 hours of each recording, different *sl* values produced different samples number and length (the lower the *sl* values, the higher the number of samples, the shorter the sample). The predictive value of hyperparameter *sl* towards validation QCK was fairly low relatively to other hyperparameters.

**Main results** Our best ANN model achieved an average QCK across HDRS and YMRS items of 0.609 in **close-to-interview** samples, a value that can be semi-qualitatively interpreted as moderate agreement. Item level QCK correlated weakly with item class imbalance but fairly with item Shannon’s entropy. Table 2 shows QCK for each item in HDRS and YMRS. Briefly, QCK was highest for *H12 somatic symptoms gastrointestinal* (0.775) and lowest for *H10 anxiety psychic* (0.492). *H10* had also the highest entropy (1.370), however, *H7 work and activities*, despite having the second highest entropy (1.213), had a QCK of 0.629, ranking as 9th best predicted item.

**Table 2:** Performance ranges from 0.775 on “somatic symptoms gastrointestinal” and to 0.492 on “anxiety psychic”. Item level performance across Hamilton Depression Rating Scale (a) and Young Mania Rating Scale (b) items. QCK: Quadratic Cohen’s *κ*.

|  |  |  |  |  |  |  |  |  |  |  |
| --- | --- | --- | --- | --- | --- | --- | --- | --- | --- | --- |
| QCK | H1<br>0.642 | H2<br>0.624 | H3<br>0.694 | H4<br>0.534 | H5<br>0.595 | H6<br>0.512 | H7<br>0.629 | H8<br>0.604 | H9<br>0.508 | H10<br>0.492 |
|  | H11<br>0.636 | H12<br>0.775 | H13<br>0.582 | H14<br>0.594 | H15<br>0.691 | H16<br>0.637 | H17<br>0.574 | Y1<br>0.602 | Y2<br>0.590 | Y3<br>0.627 |
|  | Y4<br>0.629 | Y5<br>0.591 | Y6<br>0.572 | Y7<br>0.582 | Y8<br>0.588 | Y9<br>0.755 | Y10<br>0.602 | Y11<br>0.566 |  |  |

**Table 3:**
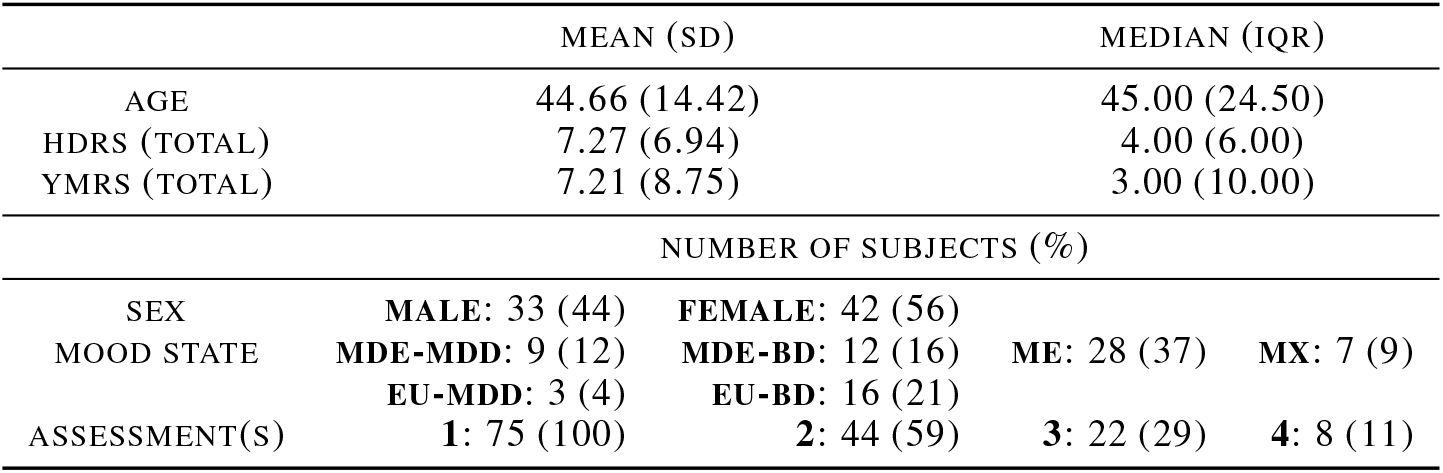
Clinical-demographic characteristics of the study population (N = 75). According to the DSM-5, a mood episode can be categorized as either a major depressive episode or a manic episode. As a bridge between these two, the DSM-5 admits a mixed symptoms specifier to cases where symptoms from both polarities are present. SD: standard deviation; IQR: inter-quartile range; MDE-MDD: Major Depressive Episode in Major Depression Disorder; MDE-BD: Major Depressive Episode in Bipolar Disorder; ME: Manic Episode; MX: Mixed symptoms episode; Eu-BD: Euthymia in Bipolar Disorder; Eu-MDD: Euthymia in Major Depressive Disorder.

**Shift over time** When tested on **far-from-interview** samples, our system overall had a drop in performance. The average QCK was 0.498, 0.303, 0.182 on segments taken respectively from the first, second, and third thirty-minute interval. After this decline the average QCK fluctuated through the following thirty-minute intervals with 0.061 as the lowest value 15 hours into the recording. The items with the biggest drop in QCK relatively to their baseline value were 1) *H9 agitation, H10 anxiety somatic, Y4 sleep*, and *Y9 disruptive-aggressive behavior* in the first thirty-minute interval 2) *H9 agitation, H10 anxiety somatic, Y9 disruptive-aggressive behavior*, and *H4 early insomnia* in the second thirty-minute interval 3) *Y4 sleep, H4 early insomnia, Y9 disruptive-aggressive behavior*, and *H10 anxiety somatic* in the third thirty-minute interval. On the other hand, items that retained their original QCK value the most were: a) *H1 depressed mood, Y11 insight, H2 feelings of guilt* in first thirty-minute interval; b) *H17 insight H1 depressed mood* and *H2 feelings of guilt* in the second thirty-minute interval; c) *H4 early insomnia, Y4 sleep, H10 anxiety somatic*, and *Y9 disruptive-aggressive behavior* in the third thirty-minute interval. This pattern matched clinical intuition as items in the former group can be relatively more volatile and more reactive to environmental factors (including medications), whereas items in the latter group tend to be change more slowly. Item level QCK across successive thirty-minute intervals is shown in Figure A.2.

### 2.3 Post-hoc diagnostics

In order to gain further insights into the errors that our system made on **close-to-interview** holdout samples, we studied distribution of residuals, i.e. the signed difference between predictions (model outputs) and ground truth (clinician scores). For the sake of better comparability, items with a rank step of two (e.g. *Y5 irritability*) were re-scaled to have a rank step of one like other items. Figure 3 illustrates that the model was correct most of the times, residuals were in general evenly distributed around zero, and when wrong the model was most often off by just one. Consistently with item level QCK values, *H10 anxiety psychic* stands out from other HDRS item as one particularly difficult to predict, with a residuals distribution that is slightly skewed towards negative values; in other words, the model tends to predict a higher score than the ground truth.

**Figure 3:**
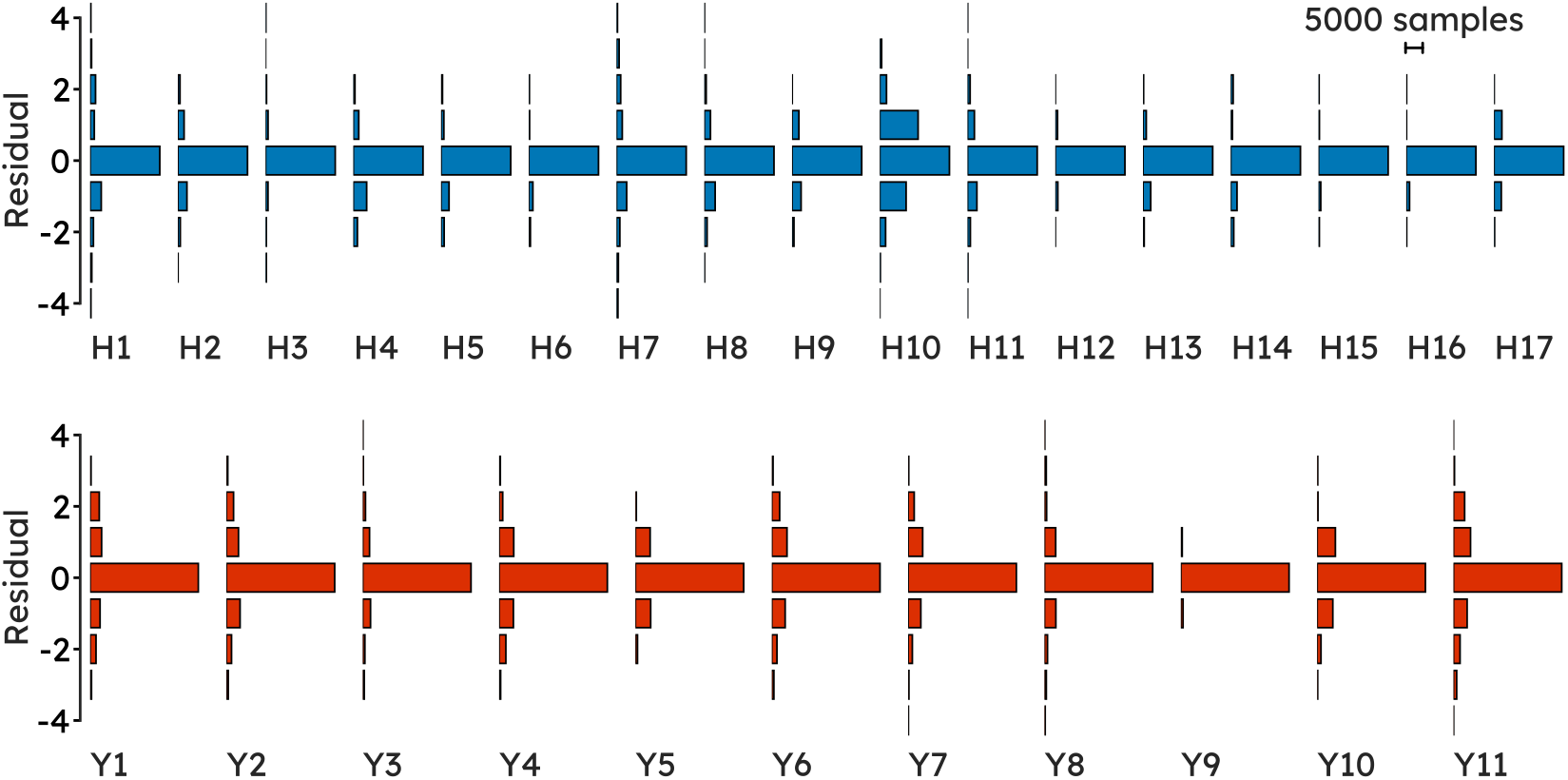
Item residuals overall show a symmetric distribution centered around zero, showing that the model is correct most of the time, it is not systematically either under- or over-predicting, and when wrong, it is usually off by only one. Residuals, signed difference between prediction (*ŷ*) and ground truth (*y*), are shown across Hamilton Depression Rating Scale (Top) and Young Mania Rating Scale (Bottom) item.

Furthermore we investigated the correlation structure among item residuals to check whether any meaningful pattern emerged. Figure 4 shows the undirected graphical model for the estimated probability distribution over HDRS and YMRS item residuals. The graph only has positive edges, that is only positive partial correlation between item residuals and co-variates. HDRS and YMRS nodes tend to have weak interactions across the two scales, with the exception of nodes that map the same symptom, e.g. *Y11* and *H17* both query *insight*. Within each scale, partial correlations are stronger among nodes underlying a common symptom domain, e.g. *H1* and *H2* constitute “core symptoms of depression” [39], and speech (*Y6*) is highly related to mood (*Y1*) and thought (*Y7, Y8*) [82]. Average node predictability for HDRS and YMRS items, a measure of how well a node can be predicted by nodes it shares an edge with, akin to *R*^2^, was 48.428%.

**Figure 4:**
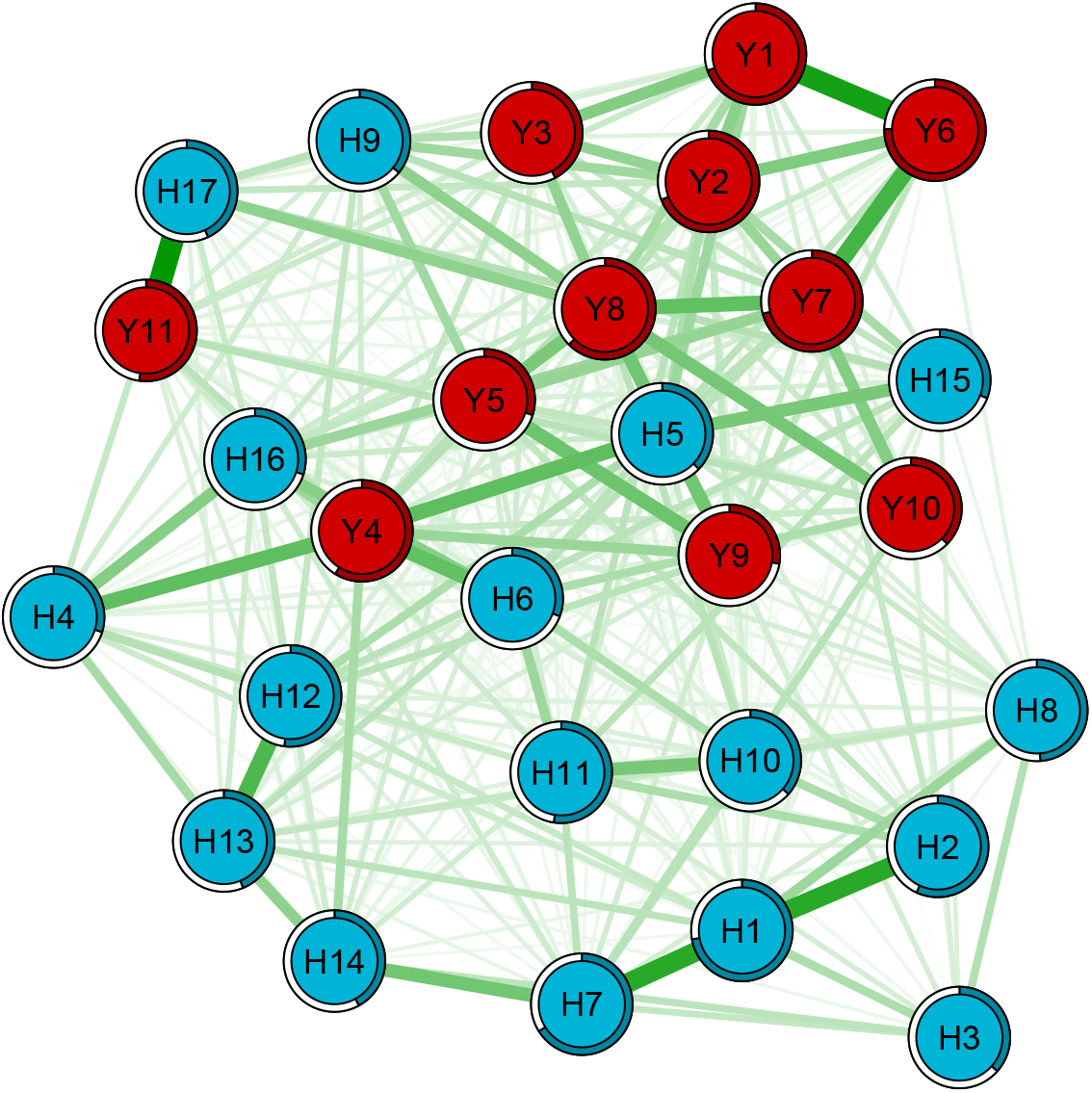
Partial correlations between item residuals indicate that the model learned the scales natural correlation structure. Network displaying the relationship between Hamilton Depression Rating Scale (blue) and Young Mania Rating Scale (red) item residuals. Green edges represent positive partial correlations between variables. Rings around nodes represent variance in a given variable with shadowed parts displaying the proportion of variance in that node that is explained by nodes that connect with it. Connections are stronger between items underpinning a common symptom domain. H: Hamilton Depression Rating Scale; Y: Hamilton Depression Rating Scale

Stability analyses in which we bootstrapped all models 500 times showed that some edges were estimated reliably (i.e. they were included in all or nearly 500 bootstrapped samples), but there also was considerable variability in the edge parameters across the bootstrapped models. Individual edges and their rank order should be interpreted with care.

### 2.4 Channels contribution

We were interested in whether physiological data modalities contributed differently towards performance across items. This question, further to clinical relevance, has also practical implications since other devices may not implement the same sensors as Empatica E4. Figure 5 shows that while all modalities seem to positively contribute to test performance across items, this is markedly the case with ACC as the model records the biggest drop in performance upon removal of this channel from input features. Specifically, upon zeroing out the contribution of ACC, the biggest deterioration in performance was observed for a) items mapping anxiety, i.e. *H12 somatic symptoms gastrointestinal* (relative difference in QCK Δ_*QCK*_ = −0.403), *H14 genital symptoms* (Δ_*QCK*_ = −0.334), *H11 anxiety somatic* (Δ_*QCK*_ = −0.321), *H13 general somatic symptoms* (Δ_*QCK*_ = −0.285), *H15 hypocondriasis* (Δ_*QCK*_ = −0.282), b) *YMRS4 sleep* and *YMRS9 disruptive-aggressive behavior* (with a Δ_*QCK*_ of −0.371 and −0.281 respectively), and c) core depression items, i.e. *H1 depressed mood* (Δ_*QCK*_ = −0.276) and *H2 feelings of guilt* (Δ_*QCK*_ = −0.263). On the other hand, the contribution of BVP was relatively modest since upon dropping this channel items generally had only a marginal reduction in QCK, most marked for *H16 loss of weight* (Δ_*QCK*_ = −0.0953) and *H11 anxiety somatic* (Δ_*QCK*_ = −0.0780).

**Figure 5:**
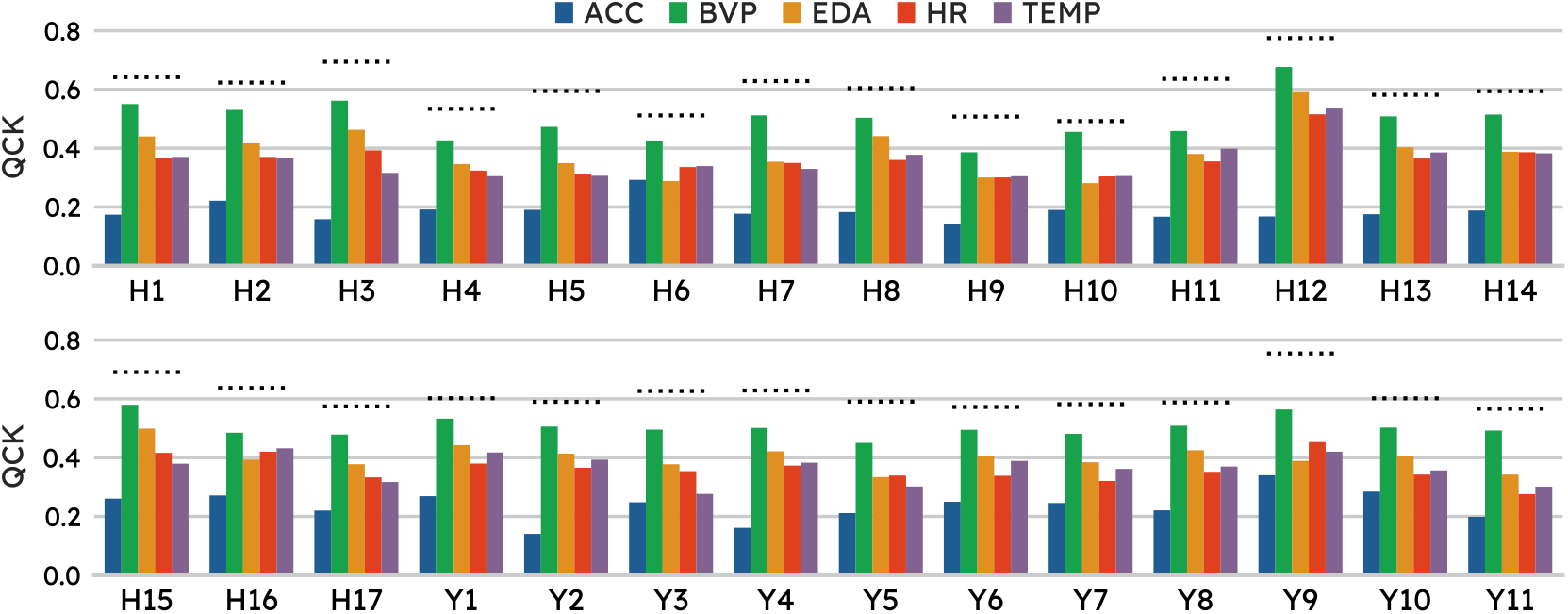
Overall all physiological data modalities contributed towards the test performance across item, however this was particularly pronounced for acceleration, whereas the model performance was only marginally impacted by the removal of blood volume pressure from input features. Effect of dropping individual channels on the performance across items. The dotted line is at the level of baseline model performance on a given item while each bar indicates the performance upon re-training the best model including all channels but the one corresponding to the bar color code, as shown in the legend. ACC: acceleration; BVP: blood volume pressure; EDA: electrodermal activity; HR: heart rate; QCK: Quadratic Cohen’s *κ*; TEMP: temperature.

## 3 Discussion

In this work we introduced a new task in mood disorders monitoring with personal sensing, that is inferring all 28 items from HDRS and YMRS, the most widely used clinician-administered scales for depression and mania respectively, the two polarities of mood disorders. While previous studies aimed to predict a single label, e.g. the disease status or the total score on a psychometric questionnaire, we advocate for modeling the full mood disorders symptom profile since such information, useful towards precision psychiatry, goes lost when reduced to a single label. This new task is complex and comes with a number of methodological challenges, some of which we have herewith analyzed.

We developed and tested our framework using samples taken in the five hours following the clinical interview (**close-to-interview** samples). Our model showed moderate agreement [51] with expert clinician (average QCK of 0.609) on holdout samples. The item level performance showed fair correlation with the item entropy, indicating that items with a higher “uncertainty” in their sample distribution tend to be more difficult to predict. Difference in entropy is partly inherent to the scale design as the number of ranks is not the same across all items. Among the explored options of segmentation window length, 16 seconds was used in the selected model. While this might be an optimal duration, it should also be noted that higher segmentation window length values translated in fewer segments available for model development. The space of explored solutions to the problem at hand included a hyper-parameter 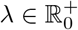, which controlled the degree to which the model should be encouraged to learn cross-subjects invariant representations when solving the main task (HDRS and YMRS inference). A *λ* value of zero means that no penalty is paid for learning to distinguish among subjects from the learned representation. Of notice, *λ* was non-zero in the optimal model, yet the training curve showed that the reduction of the main loss across epochs was paralleled by an increase in subjects identifiability from the learned representations. This suggests that an ANN (even a simple architecture such as the one we employed) tends to become reliant on representations that encode subject specific information for solving the task at hand.

When used on samples collected from thirty-minute sequences following the first five hours of the recordings (**far-from-interview** samples), our model had a significantly lower performance with average QCK declining down to 0.182 in the third half-hour and then oscillating but never recovering to the original level. Across the first three thirty-minute intervals, the items suffering the sharpest decline relatively their baseline performance were items mapping symptom involved with sleep or psycho-motor activity, i.e. *H4 early insomnia, H9 agitation, H10 anxiety somatic, Y4 sleep*, and *Y9 disruptive-aggressive behavior*. These symptoms likely have some natural variability over time, especially in response to drugs administration. On the other hand, symptoms corresponding to more stable phenomena and arguably less reactive to drugs administration in the short-term were those retaining a QCK closest to the corresponding baseline value, i.e. *H1 depressed mood, H2 feelings of guilt*, and *Y11*/*H17 insight*. While a departure from the scores collected at the start of the recording seem plausible, especially for some items, a shift in the physiological data distribution is very likely in a naturalist setting. This points to the importance of testing in samples collected at different points in time than those used for model development. Furthermore, this also highlights the methodological challenge of learning representations invariant to background (latent) features, unrelated to the target variable, that may dominate the signal and may be unduly exploited by ANNs, foiling generalization.

Residuals from predictions on holdout **close-to-interview** samples showed a symmetric distribution, centered around zero, showing that the model was not systematically predicting either higher or smaller values than the ground truth. The network of item residuals illustrated that our model erred along the correlation structure of the two symptom scales, as stronger connections were observed among items mapping the same symptom or a common domain. In other words, whenever the model mispredicted a given symptom it also tended to mispredict symptoms from the same domain. Re-training our best model including all input channels but one showed that acceleration was the most important modality. Coherently, items whose QCK deteriorated the most upon removing this channel were those mapping symptom domains clinically observable through patterns of motor behavior.

As this work proposes an novel framework to predict individual items in psychometric scales, we would like to highlight a number of limitations in our study. **(a)** Patients were scored on HDRS and YMRS by a single clinician. Having scores from multiple (independent) clinicians on the same patients would be desirable since it would help appreciate model performance in view of inter-rater agreement. We note that it was the same clinician who scored all subjects recruited for this study and thus we did not have to train multiple clinicians to reach an acceptable level of inter-rater agreement. **(b)** The lack of follow-up (independent) HDRS and YMRS scoring within the same session did not allow to estimate to what degree a shift from the baseline assessment in the items score might be at play. Relatedly, we acknowledge that the choice of five hours for our main analyses may be disputable and other choices may have been valid too; five hours is an informed attempt to trade off a reasonably high number of samples with a minimization in the distribution shift over both target variables and physiological data. On the other hand, the main point of this study was to suggest a novel task and to explore related ML challenges; thus studying the effect of different time cut-off for the main analyses was not within the scope of this work. **(c)** Given the naturalist setting, patients were on medications, which might interfere with physiological variables being recorded. **(d)** As this was a single center study, further investigations are needed using multi-center data to assess generalization to different hospitals and potentially different countries.

### Conclusion

In summary, we presented a novel task in personal sensing for mood disorders, that is inferring individual symptoms severity. We believe that this task is better aligned with the objectives of personalized medicine than other tasks suggested in previous works since it enables matching different clinical profiles with the most appropriate treatment plan. This would not be possible if the disease state or the overall severity level is all we know about a patient. We illustrated some of the associated ML challenges and explored possible approaches. The most difficult hurdle towards real-world implementations is generalization to future points in time. A departure from the scores collected during the interview seems reasonable, especially for some items and, in this sense, having more frequent observation might help. However, a shift in the distribution of physiological data is likely the major obstacle as subjects go through different patterns of activity during the day and move across different environments.

### Future directions

Results from the analyses herewith presented and related limitations point to future directions we believe are worth investigating. **(i)** The decline in performance over future points in time stands out the main challenge towards real-world implementations and suggests that the model struggles to adapt to changes in background (latent) variables, e.g. changes in activity patterns and/or environment. Research into domain adaptation should therefore be prioritized. We also speculate that mood disorders symptomatology and relevant physiological data might have slowas well as fast-changing components. A model with a segment length of 16 seconds would seem unsuitable for representing the latter. Trying to explicitly capture both dynamics is therefore something we will explore in the future. **(ii)** Generalization to patients unseen during model development is a desirable property towards real-world applications of a mood symptoms scoring system and something we consider exploring in the future. Another approach altogether to tackle this point, sidestepping considerations of cross-subjects variability, is to develop (or fine-tune) a model for each individual patient, which has been explored in other related fields [8, 61]. **(iii)** As supervised learning performance has been shown to grow roughly logarithmically in the size of annotated datasets [72], annotation remains a major bottleneck in biomedical settings because it is, as the field of EMA in mood disorders shows, notoriously resource-intensive to obtain. Self-supervised-learning approaches would seem the way forward towards real-world applications [42], as shown for example with large language models where pre-training in a self-supervised way enabled tremendous gains in performance [20, 58]. Yet, relatively little research has been done into its use in personal sensing. Lastly, **(iv)** For a ML system to be trustworthy and actionable in a clinical setting, further research into model explainability (i.e. why a given output is produced) and uncertainty is needed [23, 37].

## 4 Methodology

### 4.1 Study protocol

The analyses herewith presented rely on an original dataset being collected as part of a prospective, exploratory, observational, single-center, longitudinal study with a fully pragmatic design embedded into current real-world clinical practice. The study was conducted in compliance with the ethical principles of medical research involving humans (WMA, Declaration of Helsinki and Hospital Clinic Ethics & Research Board (HCB/2021/104, HCB/2021/1127)). Subjects with a DSM-5 diagnosis of either MDD or BD were recorded with an Empatica E4 and were scored by a clinician on HDRS and YMRS. Further details are given in Anmella et al. [4].

### 4.2 Pre-processing and data selection

The raw data from an E4 Empatica recording session comes as a collection of 1D arrays of recorded sensory modalities. We quality-controlled our data with the rules by Kleckner et al. [40] and the addition of a rule to remove HR values that exceeded the physiologically plausible range (25-250 bpm). Each quality-controlled recording session was then segmented using a sliding window, whose length *sl* (in real time seconds) is a hyperparameter, enforcing no overlap between bordering segments (to prevent models from exploiting overlapping motifs between segments). These segments and the corresponding clinician-scored psychometric questionnaires (HDRS and YMRS) from the subjects wearing the E4 device formed our dataset, 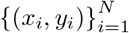, consisting of *N* observations, where *x*_*i*_ is a recording segment including different sensory modalities (i.e., ACC, BVP, EDA, HR, TEMP) and *y*_*i*_ is a 28-entry vector resulting from the HDRS and YMRS concatenation. **Close-to-interview** samples, i.e. segments from the first five hours of recordings, were split into train, validation, and test sets with a ratio of 70-15-15. Segments from the following thirty-minute intervals of recording (**far-from-interview**) were used for inference only to study how the developed model responded to distribution shift across time.

### 4.3 Evaluation Metric

HDRS and YMRS items are ordinal in nature (see Appendix B and Appendix C), that is to say although there is a finite set of possible labels like in any classification task, the labels present a natural inherent order among themselves like in regression problems: each HDRS and YMRS item admits an ordered sequence of ranks (also referred to as classes or scores) ⟨*y*_*j,k*_ = *y*_*j*,1_ ≺ … ≺ *y*_*j,k*_⟩, where *j* indexes the items/tasks and *k* indexes the item ranks. By way of example, *H11 anxiety somatic* can be scored as 0-Absent, 1-Mild, 2-Moderate, 3-Severe, or 4-Incapacitating. As expected, the item distribution was imbalanced towards low scores. This is because patients on an acute episode usually receive intensive treatment and acute states therefore tend to be relatively short-lived periods in the overall disease course [17, 79]. Metrics accounting for class imbalance should be used when evaluating a classification system in such a setting, since, otherwise, a trivial system assigning all items to a single class (e.g. the corresponding majority item rank) could outperform genuinely engineered systems.

Baccianella et al. [5] proposed the use of macro-averaged versions of common distance metrics (e.g. the mean absolute error) for the evaluation of a classifier in a setting with imbalanced ordinal data. We preferred Cohen’s *κ*, in particular its quadratic version, since, further to its suitability to imbalanced ordinal data, it is familiar and easily interpretable to clinicians [70], i.e. it expresses the degree to which the ANN learned to score segments in agreement with the clinician’s assessments. QCK penalizes disagreements proportionally to their squared distance and is defined as:

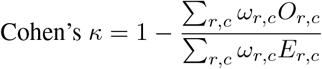

where *r, c* ∈ {1, 2, …, *K*} respectively index the rows and columns of three square matrices *ω, O*, and *E. K* is the number of classes, for instance, *K* = 5 for *H1 depressed mood* and *K* = 3 for *H4 early insomnia. O*_*r,c*_ is the number of observations, whose ground truth (clinician’s score in our use case) is the *c*^*th*^ class, that are classified in the *r*^*th*^ class by the prediction model. *E* is the outer product between the two classification histogram vectors (prediction and ground truth), normalized such that *E* and *O* have the same sum. *ω*_*i,j*_ is weight penalization for every pair *r, c*. The quadratic weightage sets *ω*_*r,c*_ = (*r* − *c*)^2^*/*(*K* − 1)^2^.

### 4.4 Model Design

The task at hand is supervised, specifically we sought to learn a mapping from the recording segment to HDRS and YMRS scores: *f* : *x*_*i*_ ↦ *ŷ* _*i*_. We used ANNs as model class for *f* (·). The model we developed comprised two independent sub-models (Figure 2):

1. a **classifier** (CF), which itself consisted of three sequential modules: (1.1) a *channel encoder* (EN) for projecting sensory modalities onto the same dimensionality, (1.2) a *representation module* (RM), for extracting features, and lastly (1.3) 28 parallel *item predictors* (IP), indexed by *j* ∈ {1, …, 28} and each learning *p* (*y*_*i,j*_ |IP_*j*_ (*h*_*i*_)), that is the conditional distribution over ranks for the *j*^*th*^ item given *h*_*i*_ = RM (EN (*x*_*i*_));
2. a **critic** (CR), using the representation from RM for telling subjects apart. The critic competed in an adversarial game against the channel encoders and the representation module, designed to encourage cross-subjects invariant representations.

#### 4.1.1 Classifier

The task of inferring HDRS and YMRS items is an instance of MTL (**c1**), to which we adopted a hard parameter-sharing approach: all tasks shared the same model trunk RM (EN (·)), and thus the same base representation of the input data *h* = RM (EN (·)), which was then distributed across task-specific layers, IP. Comparatively to developing as many independent learners as there are tasks, MTL offers advantages such as improved data efficiency, reduced overfitting through shared representations, and fast learning by leveraging auxiliary information. However, in some pathological instances, tasks might interfere with each other, a phenomenon known as negative transfer [13]. As usually done with hard parameter-sharing [60], the multi-task loss was set equal to the average of task-specific losses, each weighted by the corresponding item rank step to account for different item weights on the scale total score:

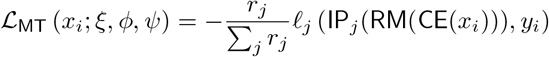

where *r*_*j*_ is the rank step size of the *j*^*th*^ item and *ξ, ϕ*, and *ψ* are respectively EN, RM, and IP parameters. Details on the specific form of *ℓ*, the task-loss, are given below.

##### Channel encoders

Since the sampling rate varies across the recorded signals within a segment, these are typically time-aligned, e.g. to the level of a second in wall-time [1, 45]. However, the down-sampling process employed to time-align data (usually via max-pooling or averaging) can risk removing useful information in the raw recordings. Thus, we took a different approach, shown to be more effective in this setting [44], whereby each channel was mapped to the same dimensionality with the use of an channel encoder. We experimented with a simple Multilayer Perceptron (MLP), a Gated Recurrent Unit (GRU) [11] or, alternatively, the Time2Vec representation proposed in [38]. We could then concatenate the encoded representations and feed them to RM in the same manner as the time-aligned data. Prior to passing segments through EN, each channel was re-scaled. The optimal embedding dimensionality and re-scaling type (either standardization or normalization) were set during tuning.

##### Representation module

A class of deep learning architectures specifically engineered to exploit dependencies in time series data are Recurrent Neural Networks (RNNs) [48]. These consume an input sequence one time-step at a time and encode historical information from previous time-steps in a hidden state so that at any given time point, further to the current time point input, the model receive information about previous time points passed on down a hidden state (a recurrence link), sequentially updated with each new time step. Specifically, we used a single-layer BiLSTM [66] for RM. This consists of two LSTMs, one taking the input in a forward direction, and the other in a backward direction, thereby improving the representation of temporal information in the model.

##### Item predictors (c2)

The extracted representation *h* = RM (EN (·)) was then used as input to 28 IP, each dedicated to a specific HDRS/YMRS item. We experimented with three different treatments of the target variables, each translating to a different set up of the task-specific layer and the task-specific loss.

1) Each item score prediction was treated as a multi-class classification problem, therefore ignoring the natural order among item ranks. Accordingly, the *j*^*th*^ item predictor consisted of a fully-connected layer, with as many output units as the number of ranks under that item, to which a softmax activation was applied and the categorical cross-entropy (CCE) was used as loss function.

2) We used the same task-specific architecture as in 1) but adopted the QWK loss, as proposed in de La Torre et al. [18], which re-writes Cohen’s *κ* in terms of probability distributions.

3) We implemented the ordinal neural network transformation model (ONTRAM) [41] which parameterize the CCE loss to incorporate the order of the outcome, by deriving class probabilities from the conditional density function of a latent variable onto which observed classes are mapped.

#### 4.4.2 Critic (c3)

We encouraged cross-subjects invariance in the representation extracted with RM (EN (·)) by adding a critic CR, whose task was to correctly distinguish subjects apart from extracted representation, in an adversarial game, similarly to [10, 55].

Concretely, the critic, a simple MLP, took as input the extracted representation *h* and was trained to identify subjects from it. CR’s task was therefore to minimize, with respect to CR’s parameters *θ*, the following CCE loss:

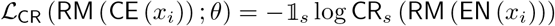

where 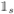 is an indicator taking value 1 when the *i*^*th*^ segment belongs to the *s*^*th*^ subject and 0 otherwise, and CR_s_() is the critic output (i.e. a probability from a softmax activation) for the *s*^*th*^ subject. On their part, EN and RM tried to trump the CR by filtering out from *h* information that could make the CR’s task easy, while, at the same time retaining enough useful information for the item predictors IP_j_. To achieve this, the following term was added to ℒ_*MT*_ (the multi-task loss), which was minimized with respect to EN’s and RM’s parameters, *ξ* and *ϕ*:

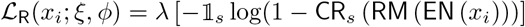

where 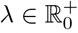. The classifier’s total loss was then ℒ_CF_ = ℒ_MT_ + ℒ_R_, where ℒ_R_ acted as a regularizer, a price the classifier CF paid for encoding subject-specific information in the representation *h* learned by RM(EN(·)). Values of *λ* trade off learning cross-subjects invariant representations *h* against solving the main objective; for *λ* = 0, no incentive is given towards learning cross-subjects invariant representation. In practice at each training step, we alternated between optimization of ℒ_CF_ and ℒ_CR_, while keeping parameters of the sub-model not being optimized (respectively, *θ* and *ξ, ϕ, ψ*) fixed. Values of *λ* in [0, 1] were used before [10, 55], so we sampled values from 𝒰 _[0,1]_ during tuning.

### 4.5 Learning from imbalanced data (c4)

In a setting with class imbalance, a classifier might struggle with predicting the minority class, usually the point of interest, since this is swamped by the abundance of instances of the majority class. We adapted to our use case three popular imbalance learning approaches, as follows:

1) Focal loss [47]: the CCE loss from each item predicor IP_*j*_ was multiplied during training by *α*_*j,k*_ (1 − *p*_*j,k*_)^*γ*^ where: *α*_*j,k*_ and *p*_*j,k*_ were respectively the inverse frequency (as estimated from the training set) and predicted probability of the ground truth *k*^*th*^ rank for the *j*^*th*^ item, *γ* was a hyperparameter (usually set to 2). *α*_*j,k*_ ∈ ℝ^+^ was a rank-wise weight that was used to increase the importance of the minority class. Easily classified examples, where *p*_*j,k*_ → 1, caused the modulating factor to approach 0 and reduced the sample’s impact on the loss. 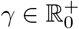 adjusted the rate at which easy examples are down-weighted.

2) Probability thresholding [9]: during inference, probabilistic predictions for each rank under the *j*^*th*^ item were divided by the corresponding rank frequency (computed on the training set). The new values were then normalized dividing each by the total sum. To avoid division by zero in the case of item ranks having zero frequency in the training set, the prior for such zero frequency ranks was set to a small non-zero value. The above is a simple *post hoc* method to re-calibrate predictions, increasing (decreasing) the predicted probability for ranks with a low (high) frequency in the training set.

3) Resampling and loss re-weighting: resampling mitigates data imbalance during training by replacing the original training set with a new one that has been resampled to be less (or not at all) imbalanced. Resampling needs some adaptation to a multi-task setting, in that resampling for one task may produce more severe imbalance across other tasks. The approach we took was to resample on the basis of the concatenation of the HDRS and YMRS severity bands: for each segment HDRS and YMRS total score was computed and binned using the cut-points suggested in [77]; each segment was therefore assigned the concatenation of such severity bands as a new label used for resampling. Thus, given a total number *B* of HDRS-YMRS severity bands in the training set, segments from each band *b* ∈ {1, .., *B*} were resampled so that their number was equal to the total number of segments divided by *B*: random under-sampling (RUS) and random over-sampling (ROS) were therefore applied to segments from relatively over-represented and under-represented bins respectively. Note that, since the same HDRS-YMRS severity bracket can be attained from multiple different combinations of HDRS and YMRS scores, the approach above mitigated but did not altogether banish unbalance at the individual item level. Furthermore, as resampling induces a departure from the original data distribution, to counterbalance this and favor learning the original data distribution, the loss of the *x*_*i*_ was rescaled proportionally to the resampling ratio of the HDRS-YMRS bin (*b*) *x*_*i*_ belonged to. That is to say, in our setting, if a segment was from a severity band on which RUS (ROS) was used, its loss (resulting from aggregation of corresponding item losses) was increased (decreased) proportionally to the resampling ratio.

#### 4.5.1 Model training and hyperparameter tuning

Given the explorative nature of this work, with various model configurations and optimization objectives, we performed an exhaustive search using the Hyperband Bayesian optimization [46] to find the hyperparameters that yield the best QCK in the validation set. All models were trained with AdamW optimizer [50] for a maximum of 400 epochs. Moreover, to speed up the training and search procedure, we employed an early stopping learning rate scheduler: we reduce the learning rate *α*_LR_ = 0.3*α*_LR_ if the model has not improved in its validation performance after 10 consecutive epochs; we terminate the training procedure if the model has not improved after 2 learning rate reductions. Dropout [71] and weight decay were added to prevent overfitting. Table 4 shows the hyperparameters search space and the configuration of the best model after 300 iterations.

**Table 4:**
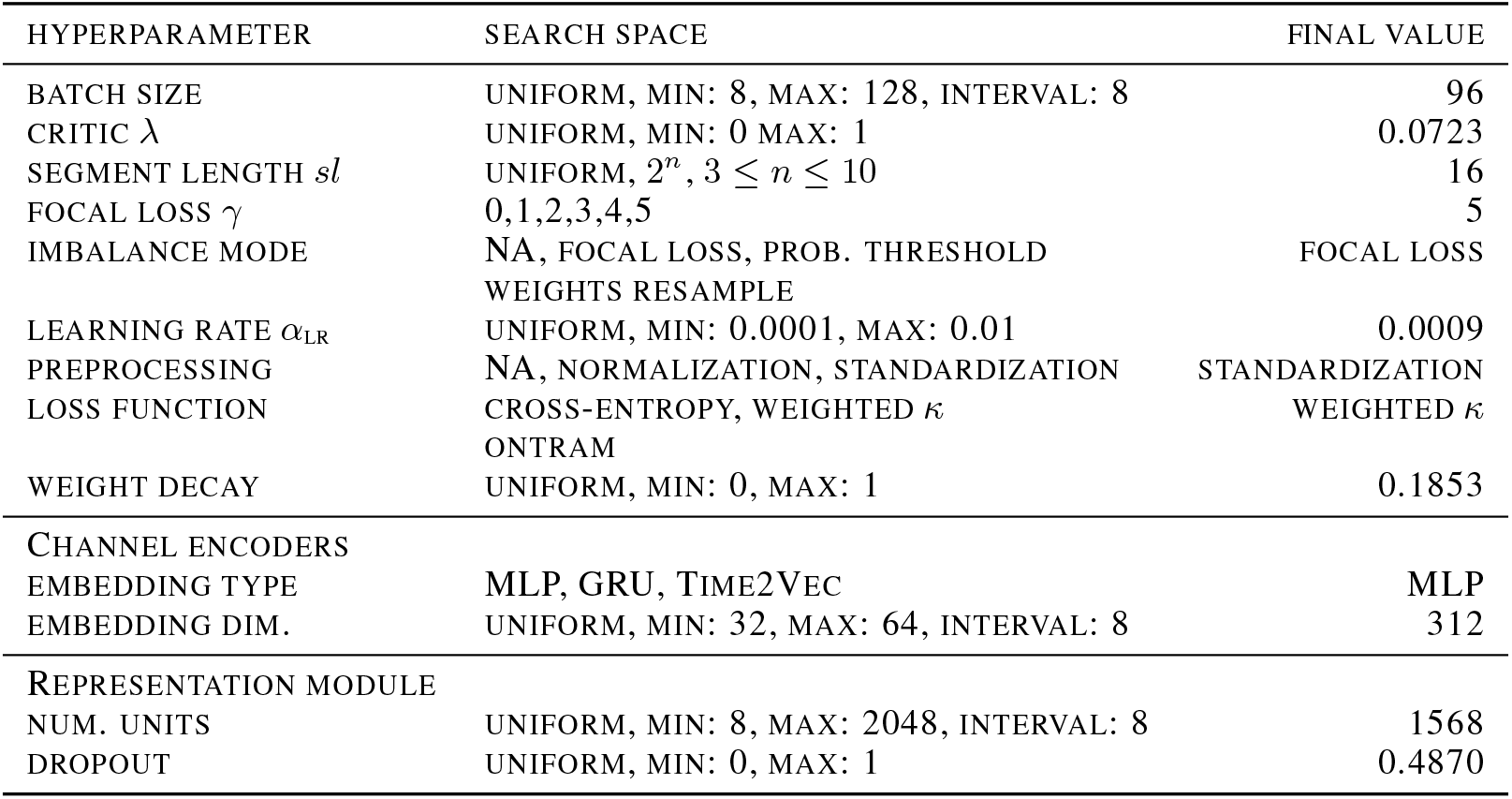
Hyperparameter search space and final configuration.

| HYPERPARAMETER | SEARCH SPACE | FINAL VALUE |
| --- | --- | --- |
| BATCH SIZE | UNIFORM, MIN: 8, MAX: 128, INTERVAL: 8 | 96 |
| CRITIC $\lambda$ | UNIFORM, MIN: 0 MAX: 1 | 0.0723 |
| SEGMENT LENGTH $sl$ | UNIFORM, $2^n$, $3 \leq n \leq 10$ | 16 |
| FOCAL LOSS $\gamma$ | 0,1,2,3,4,5 | 5 |
| IMBALANCE MODE | NA, FOCAL LOSS, PROB. THRESHOLD | FOCAL LOSS |
|  | WEIGHTS RESAMPLE |  |
| LEARNING RATE $\alpha_{LR}$ | UNIFORM, MIN: 0.0001, MAX: 0.01 | 0.0009 |
| PREPROCESSING | NA, NORMALIZATION, STANDARDIZATION | STANDARDIZATION |
| LOSS FUNCTION | CROSS-ENTROPY, WEIGHTED $\kappa$ | WEIGHTED $\kappa$ |
|  | ONTRAM |  |
| WEIGHT DECAY | UNIFORM, MIN: 0, MAX: 1 | 0.1853 |
| CHANNEL ENCODERS |  |  |
| EMBEDDING TYPE | MLP, GRU, TIME2VEC | MLP |
| EMBEDDING DIM. | UNIFORM, MIN: 32, MAX: 64, INTERVAL: 8 | 312 |
| REPRESENTATION MODULE |  |  |
| NUM. UNITS | UNIFORM, MIN: 8, MAX: 2048, INTERVAL: 8 | 1568 |
| DROPOUT | UNIFORM, MIN: 0, MAX: 1 | 0.4870 |

### 4.6 Prediction Error Examination

Using the best performing setting among those explored in the experiments detailed above we computed residuals (i.e. signed difference between prediction and ground truth) on **close-to-interview** samples and illustrated their distribution across items. Furthermore, towards investigating correlations between residuals, checking for any remarkable pattern in view of the natural correlation structure of HDRS and YMRS, we estimated a regularized partial correlation network, in particular a Gaussian graphical lasso (glasso) [29], over the item residuals. Network edge sparsity, to avoid false positives, is enforced with the the least absolute shrinkage and selection operator (LASSO) [75], which indeed shrinks all edge-weights towards zero and sets small weights to exactly zero. The strength of the regularization is traded-off by a hyper-parameter *λ*, selected with the Extended Bayesian Information Criterion (EBIC) [28]. The EBIC itself has a tuning parameter *γ*, controlling the trade-off between sensitivity and precision, which, as in Haslbeck and Waldorp [33], we set to 0.25. We also estimated node predictability, measuring how well a node can be predicted by nodes it shares an edge with, which can be interpreted similarly to *R*^2^ [34]. Lastly, bootstrapping routines were used to gain insight into the stability of the estimated parameters.

### 4.7 Channels Importance

We took a simple, model-agnostic approach to assessing each individual channel contribution to the task at hand. That is to say, we selected the system performing best on the task and re-trained it including all channels (tri-axial ACC, EDA, BVP, HR, and TEMP) but one. For each left-out channel we measured the difference in performance across items relatively to the baseline model (the one trained on all channels).

## Data Availability

Data in de-identified form may be made available from the corresponding author upon reasonable
request.

## Code availability

The codebase developed for this work is available at repository to be released upon acceptance for publication. Python 3.10 programming language was used for the symptoms scoring system, where deep learning models were implemented in PyTorch [56] while hyperparameter tuning and visualization model performance were performed in Weight & Biases [7]. All models were trained on a single Nvidia RTX 2080Ti GPU. Graphical modeling of the residuals and related analyses were performed in R 4.2.2 using packages *qgraph* [26] for network estimation and visualization, and *bootnet* [25] for bootstrapping.

## Data availability

Data in de-identified form may be made available from the corresponding author upon reasonable request.

## Acknowledgments

We acknowledge the contribution of all the participants of the study.

We thank Dr Arno Onken for reviewing the manuscript and providing insightful suggestions to improve this work.

F.C. and B.M.L. are supported by the United Kingdom Research and Innovation (grant EP/S02431X/1), UKRI Centre for Doctoral Training in Biomedical AI at the University of Edinburgh, School of Informatics. For the purpose of open access, the author has applied a creative commons attribution (CC BY) licence to any author accepted manuscript version arising.

G.A. is supported by a Rio Hortega 2021 grant (CM21/00017) from the Spanish Ministry of Health financed by the Instituto de Salud Carlos III (ISCIII) and co-financed by the Fondo Social Europeo Plus (FSE+).

E.V. thanks the support of the Spanish Ministry of Science, Innovation and Universities (PI15/00283, PI18/00805, PI19/00394, CPII19/00009) integrated into the Plan Nacional de I+D+I and co-financed by the Instituto de Salud Carlos III (ISCIII)-Subdirección General de Evaluación and the Fondo Europeo de Desarrollo Regional (FEDER); the ISCIII; the CIBER of Mental Health (CIBERSAM); the Secretaria d’Universitats i Recerca del Departament d’Economia i Coneixement (2017 SGR 1365), and the CERCA Programme / Generalitat de Catalunya. We would like to thank the Departament de Salut de la Generalitat de Catalunya for the PERIS grant SLT006/17/00357.

D.H.M. is supported by a Juan Rodés JR18/00021 granted by the Instituto de Salud Carlos III (ISCIII).

## Authors contributions

F.C. and B.M.L. conceived of the study, proposed the methodology, led the development of the software codebase for the analyses, and prepared the manuscript. G.A. contributed to data collection and critically reviewed the manuscript, A.M. and M.S contributed to data collection and curated the dataset. E.V., S.M., H.W. critically reviewed the manuscript. D.H.M. is the responsible for the overall project this study relies on for data, acquired funding, and critically reviewed the manuscript. A.V. supervised and contributed to the study design, methodology development, and manuscript writing. Members of the INTRPIBD Group contributed to data collection.

## Competing interests

G.A. has received CME-related honoraria, or consulting fees from Janssen-Cilag, Lundbeck, Lundbeck/Otsuka, and Angelini, with no financial or other relationship relevant to the subject of this article.

E.V. has received research support from or served as consultant, adviser or speaker for AB-Biotics, Abbott, Abbvie, Adamed, Angelini, Biogen, Celon, Dainippon Sumitomo Pharma, Ferrer, Gedeon

Richter, GH Research, Glaxo SmithKline, Janssen, Lundbeck, Organon, Otsuka, Rovi, Sage pharmaceuticals, Sanofi-Aventis, Shire, Sunovion, Takeda, and Viatris, out of the submitted work.

D.H.M. has received CME-related honoraria and served as consultant for Abbott, Angelini, Ethypharm Digital Therapy and Janssen-Cilag with no financial or other relationship relevant to the subject of this article.

All other authors report no financial or other relationship relevant to the subject of this article.

## A Appendix

**Figure A1:**
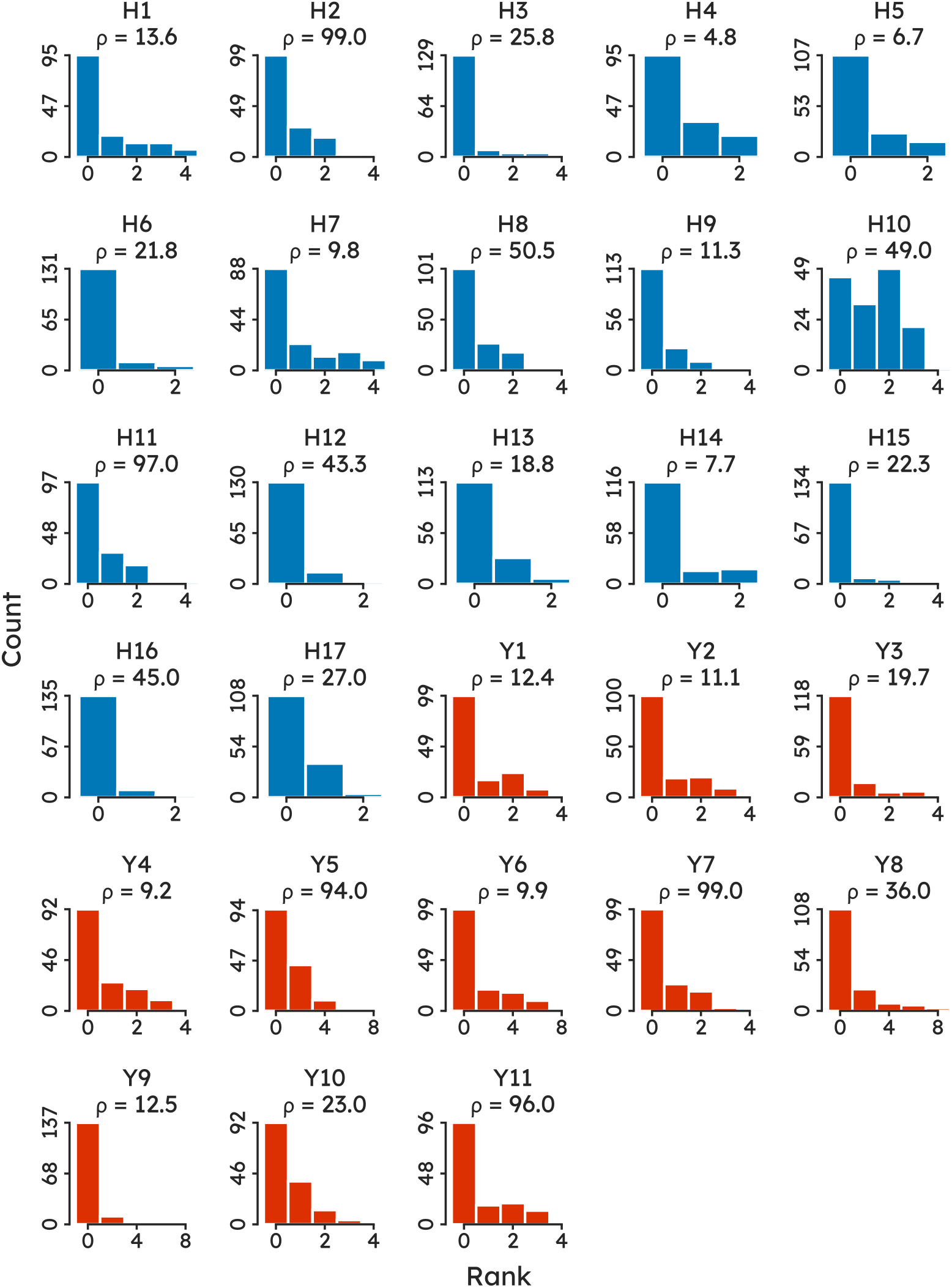
Distribution over Hamilton Depression Rating Scale (blue) and Young Mania Rating Scale (red) items across the recording sessions used in this study. The number above each barplot, *ρ*, is the cardinality of the majority class over that of the minority rank. Higher values of *ρ* thus indicate a more pronounced imbalance between the majority and the minority rank.

**Figure A2:**
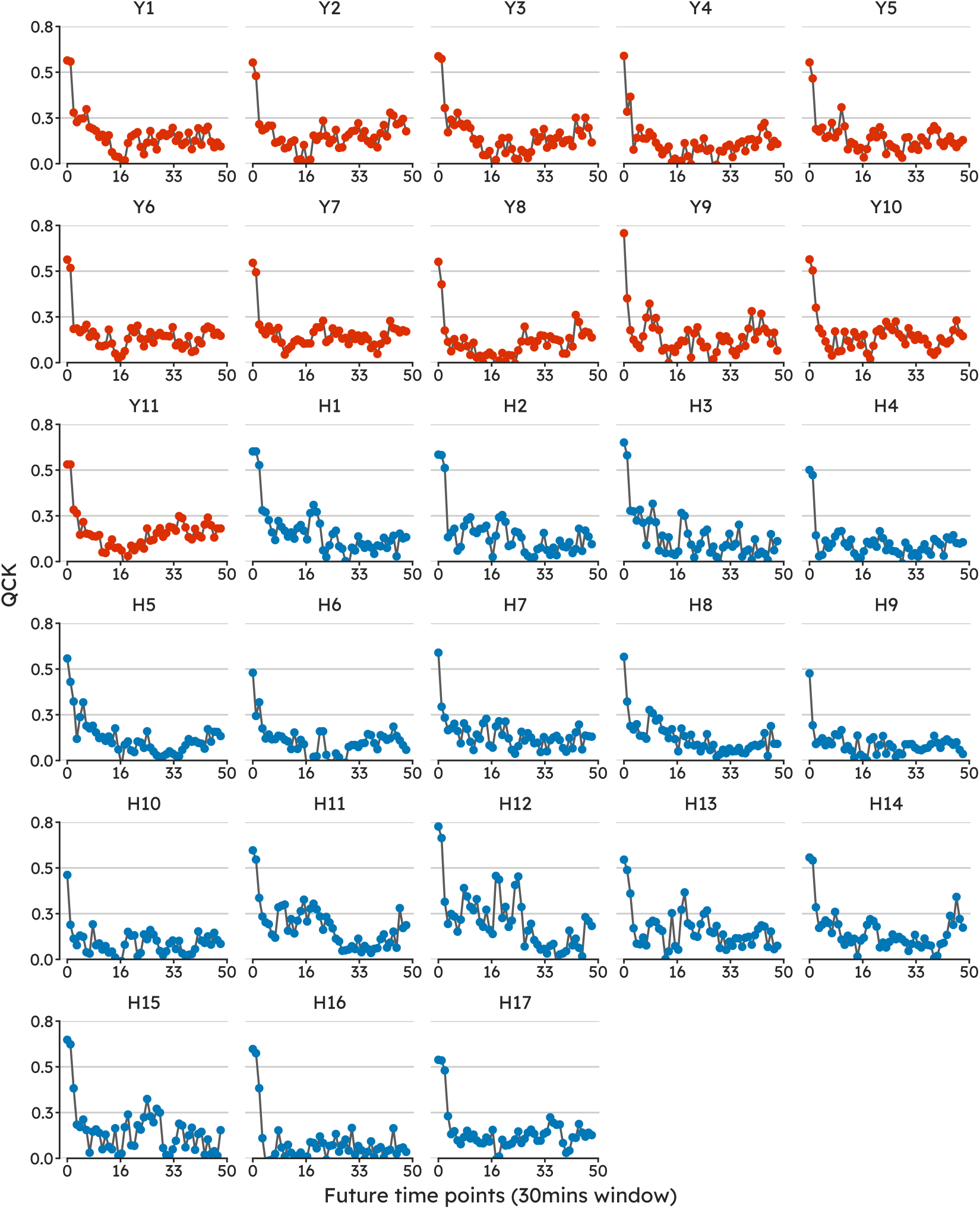
Quadratic Cohen’s *κ* (QCK) deteriorated across all items when the model was tested on segments taken further away from when the interview took place. The first point (0 on the x-axis) is the baseline performance, i.e. holdout segments from the first five hours of recordings (**close-to-interview**). Following points refer to the successive thirty minute intervals. (**close-to-interview**).

**Table A1:** Hyperparameters sorted by their importance towards predicting the monitored metric, validation Quadratic Cohen’s *κ* (QCK). QCK predictability from hyperparameters is derived by training a random forest with the hyperparameters as inputs and the metric as the target output and estimating feature importance values for the random forest. Details at docs.wandb.ai/parameterimportance.

| HYPERPARAMETER | IMPORTANCE |
| --- | --- |
| LOSS FUNCTION | 0.205 |
| DROPOUT | 0.073 |
| BATCH SIZE | 0.070 |
| WEIGHT DECAY | 0.055 |
| PREPROCESSING | 0.035 |
| NUM. UNITS | 0.033 |
| EMBEDDING DIM. | 0.032 |
| CRITIC $\lambda$ | 0.026 |
| LEARNING RATE $\alpha_{LR}$ | 0.025 |
| IMBALANCE MODE | 0.022 |
| FOCAL LOSS $\gamma$ | 0.019 |
| EMBEDDING TYPE | 0.015 |
| SEGMENT LENGTH $sl$ | 0.013 |

## B Hamilton Depression Rating Scale

Hamilton Depression Rating Scale (HDRS) [32] which consist of 17 items, each with a score to indicate severity of the symptom.

H1. Depressed Mood (sadness, hopeless, helpless, worthless)

0 - Absent.

1 - These feeling states indicated only on questioning.

2 - These feeling states spontaneously reported verbally.

3 - Communicates feeling states non-verbally, i.e. through facial expression, posture, voice and tendency to weep.

4 - Patient reports virtually only these feeling states in his/her spontaneous verbal and non-verbal communication

H2. Feelings of guilt

0 - Absent.

1 - Self reproach, feels he/she has let people down.

2 - Ideas of guilt or rumination over past errors or sinful deeds. 3 - Present illness is a punishment. Delusions of guilt.

4 - Hears accusatory or denunciatory voices and/or experiences threatening visual hallucinations.

H3. Suicide

0 - Absent.

1- Feels life is not worth living.

2 - Wishes he/she were dead or any thoughts of possible death to self. 3 - Ideas or gestures of suicide.

3 - Attempts at suicide (any serious attempt rate 4).

H4. Insomnia: early in the night

- No difficulty falling asleep.

1 - Complains of occasional difficulty falling asleep, i.e. more than 1/2 hour. 2 - Complains of nightly difficulty falling asleep.

H5. Insomnia: middle of the night 0 - No difficulty.

1 - Patient complains of being restless and disturbed during the night.

2 - Waking during the night – any getting out of bed rates 2 (except for purposes of voiding).

H6. Insomnia: early hours of the morning 0 - No difficulty.

1 - Waking in early hours of the morning but goes back to sleep.

2 - Unable to fall asleep again if he/she gets out of bed.

H7. Work and Activities 0 - No difficulty.

1 - Thoughts and feelings of incapacity, fatigue or weakness related to activities, work or hobbies.

2 - Loss of interest in activity, hobbies or work – either directly reported by the patient or indirect in listlessness, indecision and vacillation (feels he/she has to push self to work or activities).

3 - Decrease in actual time spent in activities or decrease in productivity. Rate 3 if the patient does not spend at least three hours a day in activities (job or hobbies) excluding routine chores.

4 - Stopped working because of present illness. Rate 4 if patient engages in no activities except routine chores, or if patient fails to perform routine chores unassisted.

H8. Retardation

1 - Normal speech and thought.

2 - Slight retardation during the interview.

3 - Obvious retardation during the interview. 3 - Interview difficult.

4 - Complete stupor.

H9. Agitation

0 - None.

1- Fidgetiness.

2 - Playing with hands, hair, etc.

3 - Moving about, can’t sit still.

4 - Hand wringing, nail biting, hair-pulling, biting of lips.

H10. Anxiety Psychic

0 - No difficulty.

1- Subjective tension and irritability.

2 - Worrying about minor matters.

3 - Apprehensive attitude apparent in face or speech.

4 - Fears expressed without questioning.

H11. Anxiety Somatic (physiological concomitants of anxiety)

0 - Absent.

1 - Mild.

2 - Moderate.

3 - Severe.

3 - Incapacitating.

H12. Somatic Symptoms Gastro-Intestinal 0 - None.

1 - Loss of appetite but eating without staff encouragement. Heavy feelings in abdomen.

2 - Difficulty eating without staff urging. Requests or requires laxatives or medication for bowels or medication for gastro-intestinal symptoms.

H13. General Somatic Symptoms 0 - None.

1 - Heaviness in limbs, back or head. Backaches, headaches, muscle aches. Loss of energy and fatigability.

2 - Any clear-cut symptom rates 2.

H14. Genital Symptoms 0 - Absent.

1 - Mild.

2 - Severe.

H15. Hypocondriasis

0 - Not present.

1 - Self-absorption (bodily).

2 - Preoccupation with health.

3 - Frequent complaints, requests for help, etc.

4 - Hypochondriacal delusions.

H16. Loss of Weight

0 - Less than 1 lb weight loss in week.

1 - Greater than 1 lb weight loss in week.

2 - Greater than 2 lb weight loss in week.

H17. Insight

0 - Acknowledges being depressed and ill.

1 - Acknowledges illness but attributes cause to bad food, climate, overwork, virus, need for rest, etc.

2 - Denies being ill at all.

## C Young Mania Rating Scale

Young Mania Rating Scale (YMRS) [83] which consist of 11 items, each with a score to indicate severity of the symptom.

Y1. Elevated Mood

0 - Absent.

1 - Mildly or possibly increased on questioning.

3 - Definite subjective elevation; optimistic, self-confident; cheerful; appropriate to content. 3 - Elevated; inappropriate to content; humorous.

4 - Euphoric; inappropriate laughter; singing.

Y2. Increased Motor Activity-Energy 0 - Absent.

0 - Subjectively increased.

2- Animated; gestures increased.

3 - Excessive energy; hyperactive at times; restless (can be calmed).

4 - Motor excitement; continuous hyperactivity (cannot be calmed).

Y3. Sexual Interest

0 - Normal; not increased.

1 - Mildly or possibly increased.

2 - Definite subjective increase on questioning.

3 - Spontaneous sexual content; elaborates on sexual matters; hypersexual by self-report.

4 - Overt sexual acts (toward patients, staff, or interviewer).

Y4. Sleep

0 - Reports no decrease in sleep.

1 - Sleeping less than normal amount by up to one hour.

2 - Sleeping less than normal by more than one hour.

3 - Reports decreased need for sleep. 4 - Denies need for sleep.

Y5. Irritability

0 - Absent.

2 - Subjectively increased.

4 - Irritable at times during interview; recent episodes of anger or annoyance on ward.

6 - Frequently irritable during interview; short, curt throughout.

8 - Hostile, uncooperative; interview impossible.

Y6. Speech (Rate and Amount) 0 - No increase.

2 - Feels talkative.

4 - Increased rate or amount at times, verbose at times.

6 - Push; consistently increased rate and amount; difficult to interrupt. 8 - Pressured; uninterruptible, continuous speech.

Y7. Language-Thought Disorder

0 - Absent.

1 - Circumstantial; mild distractibility; quick thoughts.

2 - Distractible, loses goal of thought; changes topics frequently; racing thoughts.

3 - Flight of ideas; tangentiality; difficult to follow; rhyming, echolalia.

4 - Incoherent; communication impossible.

Y8. Content

0 - Normal.

1 - Questionable plans, new interests.

4 - Special project(s); hyper-religious.

6 - Grandiose or paranoid ideas; ideas of reference.

8 - Delusions; hallucinations.

Y9. Disruptive-Aggressive Behavior

0 - Absent, cooperative.

2 - Sarcastic; loud at times, guarded.

4 - Demanding; threats on ward.

6 - Threatens interviewer; shouting; interview difficult.

8 - Assaultive; destructive; interview impossible.

Y10. Appearance

0 - Appropriate dress and grooming.

1 - Minimally unkempt.

2 - Poorly groomed; moderately disheveled; overdressed.

3 - Disheveled; partly clothed; garish make-up.

4 - Completely unkempt; decorated; bizarre garb.

Y11. Insight

0 - Present; admits illness; agrees with need for treatment.

1 - Possibly ill.

2 - Admits behavior change, but denies illness.

3 - Admits possible change in behavior, but denies illness.

4 - Denies any behavior change.

